# Cohort profile: The Camden Study – a pregnancy cohort study of pregnancy complications and birth outcomes in Camden, New Jersey, USA

**DOI:** 10.1101/2024.09.13.24313648

**Authors:** Stephanie Shiau, Xinhua Chen, Ayana April-Sanders, Ellen C. Francis, Shristi Rawal, Megan Hansel, Kehinde Adeyemi, Zorimar Rivera-Núñez, Emily S. Barrett

**Author notes:** **Corresponding Author:** Stephanie Shiau, PhD, MPH, Rutgers School of Public Health, 683 Hoes Lane West Office 221, Piscataway, NJ 08854 USA. These authors contributed equally (Zorimar Rivera-Núñez, Emily S. Barrett).

## Abstract

**Background:** Pregnancy is a unique stage of the life course characterized by trade-offs between the nutritional, immune, and metabolic needs of the mother and fetus. The Camden Study was originally initiated to examine nutritional status, growth, and birth outcomes in adolescent pregnancies and expanded to study dietary and molecular predictors of pregnancy complications and birth outcomes in young women.

**Methods:** From 1985-2006, 4765 pregnant participants aged 12 years and older were recruited from Camden, NJ, one of the poorest cities in the U.S. The cohort reflects a population under-represented in perinatal cohort studies (45% Hispanic, 38% non-Hispanic Black, 17% White participants; 98% using Medicaid in pregnancy). Study visits, including questionnaires, dietary assessments, and biospecimen collection, occurred in early and late pregnancy as well as at delivery. Medical records were abstracted, and a subset of mothers and infants participated in a six-week postpartum visit.

**Results:** Over the last five decades, the Camden Study has provided data toward the publication of numerous peer-reviewed papers. Results show that adolescent linear growth in pregnancy is associated with smaller birth size, possibly due to impaired hemodynamics. In the context of preterm birth and other perinatal outcomes, analyses of nutritional data have demonstrated the importance of micronutrients (e.g., folate, iron, zinc), as well as glucose/insulin dynamics and prenatal supplement use. More recent analyses have begun to unpack the biochemical pathways in pregnancy that may be shaped by race as an indicator for systemic racism.

**Conclusions:** The Camden Study data and biorepositories are well-positioned to support future research aimed at better understanding perinatal health in under-represented women and infants. Linkages to subsequent health and administrative records and the potential for recontacting participants over 18-39 years after initial participation may provide key insights into the trajectories of maternal and child health across the life course.

## Introduction

Pregnancy is a unique stage of the life course during which complications can have life-long impacts on maternal and fetal health.^1^ Even in the absence of pre-existing health conditions, a poor physiological response to the fetus’ dynamic nutritional, immune, and metabolic needs during gestation can increase the risk of perinatal complications such as preeclampsia/eclampsia, gestational diabetes (GDM), and preterm delivery.^2,3,3^ In turn, these perinatal complications increase risk of offspring developmental delay and chronic disease for both mother and offspring.^4^ In the United States (US), the prevalence of perinatal complications ranges depending on many factors. These include metabolic heterogeneity, body composition, lifestyle behaviors, as well as sociodemographic factors such as race/ethnicity that capture differences in societal and environmental influences, such as experiences of racism, poverty, social support, and education.^5–7^

The prevalence of perinatal complications as well as their associated risk factors are greater among women of color.^8–12^ For example, the rate and risk of pre-eclampsia and preterm birth are higher among Black women than other racial/ethnic groups,^11,13–15^ and Hispanic and Asian women have a 20-100% greater risk of GDM than White women.^16,17^ Although adolescent pregnancies have decreased since the 1990s, racial/ethnic and geographic disparities in adolescent birth rates remain.^18,19^ Many societal and environmental influences that increase risk of perinatal complications, such as family poverty, reduced access to community resources, and lower maternal educational attainment, are more prevalent among adolescents who become pregnant.^20^ Continued research, awareness, and attention to these racial/ethnic disparities are imperative to create health policies for communities at the most significant risk.

The Camden Study, established in 1985, was designed to address these gaps by recruiting young pregnant women from health clinics in Camden, New Jersey, one of the poorest cities in the US. The Camden Study has been funded through several major research grants that have informed the design of the study and activities. Originally, the study was initiated to examine nutritional status, growth, and birth outcomes in adolescent pregnancies (R01HD018269/Scholl; R01ES007437/Scholl) and was subsequently expanded to investigate associations of diet and biochemical markers with perinatal outcomes among young women (R01HD038329/Scholl; R21DK078865/Chen; R21HD058128/Scholl; R21HD061763/Chen R01MD007828/Chen). In this paper, we describe the design, data collection protocols, biospecimens collected, and biochemical markers measured in this cohort. We present a summary of the major findings and contributions to the field and detail future directions for research.

## Cohort description

### Study setting and recruitment

The Camden Study recruited pregnant women^a^ who received obstetric care between 1985–2006 at Cooper University Hospital, the Osborn Family Health Center, Our Lady of Lourdes Medical Center, and St. John the Baptist prenatal clinics, all located in Camden, NJ. At the time of recruitment, Camden was (and continues to be) one of the poorest cities in the United States.^21^ Initial analyses focused on differences in growth and development between adolescents and adults that potentially affect pregnancy outcomes and as such, initial study participants included primigravid and multigravid teenagers (<19 years) with a first pregnancy occurring prior to age 16, as well as older pregnant participants, aged 19-29 at first pregnancy. Subsequent analyses focused on the study of nutrition biomarkers and oxidative stress. Women with major non-obstetric health issues (e.g., non-insulin-dependent diabetes, chronic hypertension, seizure disorders, known drug or alcohol abuse issues, malignancies, leukemia, major psychiatric disorders, and lupus) at the time of recruitment were excluded.

The Institutional Review Board at Rutgers University gave ethical approval for this work. All participants signed written informed consent prior to engaging in any study activities. Data were accessed to create tables for this analysis on August 4, 2023.

### Overview of Camden Study activities

Study activities varied modestly across waves of recruitment and typically included one in-person visit in early/mid pregnancy (visit 1) followed by a subsequent visit in late pregnancy (visit 2), birth biospecimen collection, and a single in-person, postnatal visit at 4-6 weeks postpartum and up to 6 months postpartum in ∼600 participants. Biospecimens (e.g. serum, urine, saliva) were collected from participants and are stored in the Camden Study biorepository. An overview of collected measures and biospecimens/biomarkers can be found in **Tables 1 and 2**.

**Table 1.** Measures collected in the Camden Study.

| Domain | Key measures | Timepoint(s) |  |
| --- | --- | --- | --- |
|  |  | Prenatal | Postpartum |
| Maternal interviews | Sociodemographic characteristics | X | X |
|  | Reproductive and general medical history | X | X |
|  | Medications | X |  |
|  | Smoking, alcohol, and drug use | X | X |
|  | Infections | X | X |
|  | Breastfeeding |  | X |
| Maternal dietary intake | Total caloric intake | X | X |
|  | Macro- and micronutrient intake | X | X |
|  | Fatty acid intake | X |  |
|  | Glycemic index | X |  |
| Maternal anthropometric measures | Pre-pregnancy weight (recalled) | X |  |
|  | Height, weight | X | X |
|  | Skinfold thickness | X | X |
|  | Growth by knee height | X | X |
|  | Blood pressure | X | X |
|  | Gestational weight gain | X |  |
| Bone mass and density* | Dual energy x-ray densitometry (DEXA) scan | X | X |
|  | Bone ultrasound | X | X |
| Medical record review | Reproductive and general medical history | X |  |
|  | Current and prior pregnancy complications | X |  |
|  | Medications | X |  |
|  | Lab results | X | X |
|  | Delivery and neonatal information |  | X |

\* Only available in a subset of participants.

### Questionnaires

At visits 1 and 2, trained study staff conducted structured interviews to collect data on sociodemographic and lifestyle factors. These interviews included items on race/ethnicity, education, occupation, insurance status, household composition, lifestyle (e.g,. cigarette smoking, alcohol and drug use), reproductive and medical history, and family health history.

### Dietary assessments

A registered dietitian collected data on maternal diet (including use of prenatal vitamins and other dietary supplements) through 24-hour recalls administered at visits 1 and 2. These data were processed using the Campbell Master Nutrient Data Base at the Campbell Institute of Research and Technology. The nutrient intake database was updated from the United States Department of Agriculture Nutrient Database for Standard Reference and the Survey Nutrient Database for Continuing Survey of Food Intakes by Individuals as well as data from the scientific literature.^22,23^ Detailed descriptions of the dietary assessments as well as the reliability of the Camden Study’s dietary recall data have been published elsewhere.^24^ To complement the 24-hour recalls, at prenatal visits, data were collected on pre-conception vitamin and supplement use, nausea and hyperemesis, pica, food allergies, hunger, eating habits, and fasting/dieting behaviors.

### Maternal anthropometric measures

At both visits and at delivery, participants were weighed on a clinical beam-balance scale and participants reported their pre-pregnancy weight. Early pregnancy height was measured using a standard clinical stadiometer. To evaluate growth (particularly with respect to adolescent pregnancy), at study entry and again at the postpartum visit, a knee height measuring device (KHMD) was used to non-invasively measure the length of the mother’s lower leg (with 0.5 mm resolution) with four replicates collected.^25^ Knee height is a preferred measure of linear growth (particularly during pregnancy) compared to statural growth, which tends to decline during pregnancy due to vertebral compression and lordosis. A change in knee height of ≥1 mm from early pregnancy to postpartum was considered evidence of maternal linear growth during pregnancy.^26^ In a subset of participants, bone ultrasounds (Sahara, Hologic, Inc., Bedford, MA) were used to measure bone mass early/mid pregnancy and 4-6 weeks postpartum.^27^ In the same subset, bone mineral density (BMD) of the femoral neck and lumbar spine was measured by dual x-ray densitometry (DEXA) (Lunar Corp., Madison, WI; DPX-L, analysis software version 1.3y) within 2 days of delivery.^27^

In addition, at both visits, arm circumference, waist circumference, hip circumference, and thigh girth were measured by trained examiners using a standard measuring tape. To quantify adiposity, a series of skinfold thickness measurements were collected at multiple sites including subscapular, suprailiac, biceps, triceps, suprapatellar, thigh, calf, and ankle using standard protocols.^28^ All skinfold thickness measurements were taken in duplicate and averaged. Upper arm muscle area and upper arm fat area were subsequently calculated using a standard equation.^29^ Based on anthropometric measures, changes in height, weight, BMI (kg/m^2^), upper arm muscle area (cm^2^), upper arm fat area (cm^2^), and skinfold thicknesses were calculated. Finally, diastolic and systolic blood pressure were measured at both visits.

### Medical record review

After delivery, trained study personnel abstracted data from the prenatal medical record, delivery record, delivery logbooks and infant medical records. Data of interest included outcomes of all prior pregnancies, medication use, infections, fetal biometry (as determined by ultrasound), detailed data on labor and delivery for the index pregnancy, and any maternal or infant health issues. Gestational age at delivery was determined based on the last menstrual period (LMP) with confirmation or modification based on an early pregnancy ultrasound. In the case of size-for-dates discrepancies or if the LMP was uncertain, dating based on the early pregnancy ultrasound was used. Per convention, small-for-gestational age was defined as birth weight below the 10^th^ percentile and large-for-gestational age was defined as birth weight above the 90^th^ percentile-for-gestational age, adjusted for maternal parity, race/ethnicity and fetal sex.^30^ Low birth weight was defined as birth weight <2500 grams, macrosomia was defined as birth weight >4000 grams, and preterm delivery was defined as delivery <37 weeks gestation.

### Prenatal and birth biospecimen collection

At each study visit, biospecimens were collected as described briefly below. Maternal blood was collected by venipuncture with time of sample, time of last meal, and fasting status (12+ hours since last meal) recorded. Blood samples were processed and resulting aliquots of serum, whole blood, and plasma were stored at –80C pending analysis. Numerous analyses based on maternal blood samples have been conducted (**Table 2**) and additional aliquots remain in the biorepository for future analysis. In addition, urine and saliva were collected in each trimester. Urinalysis was conducted in clinical laboratories to test for medical conditions (e.g., proteinuria, kidney stones) as well as microbes and viral infections (e.g., yeast, bacterial vaginosis, enterococcus faecalis, proteus mirabilis) after which samples were frozen at −80C for future analyses. At delivery, a cord blood specimen was obtained in a subset of participants and processed with aliquots of serum, plasma, and whole blood stored in the Camden Study repository for future analysis.

**Table 2.**
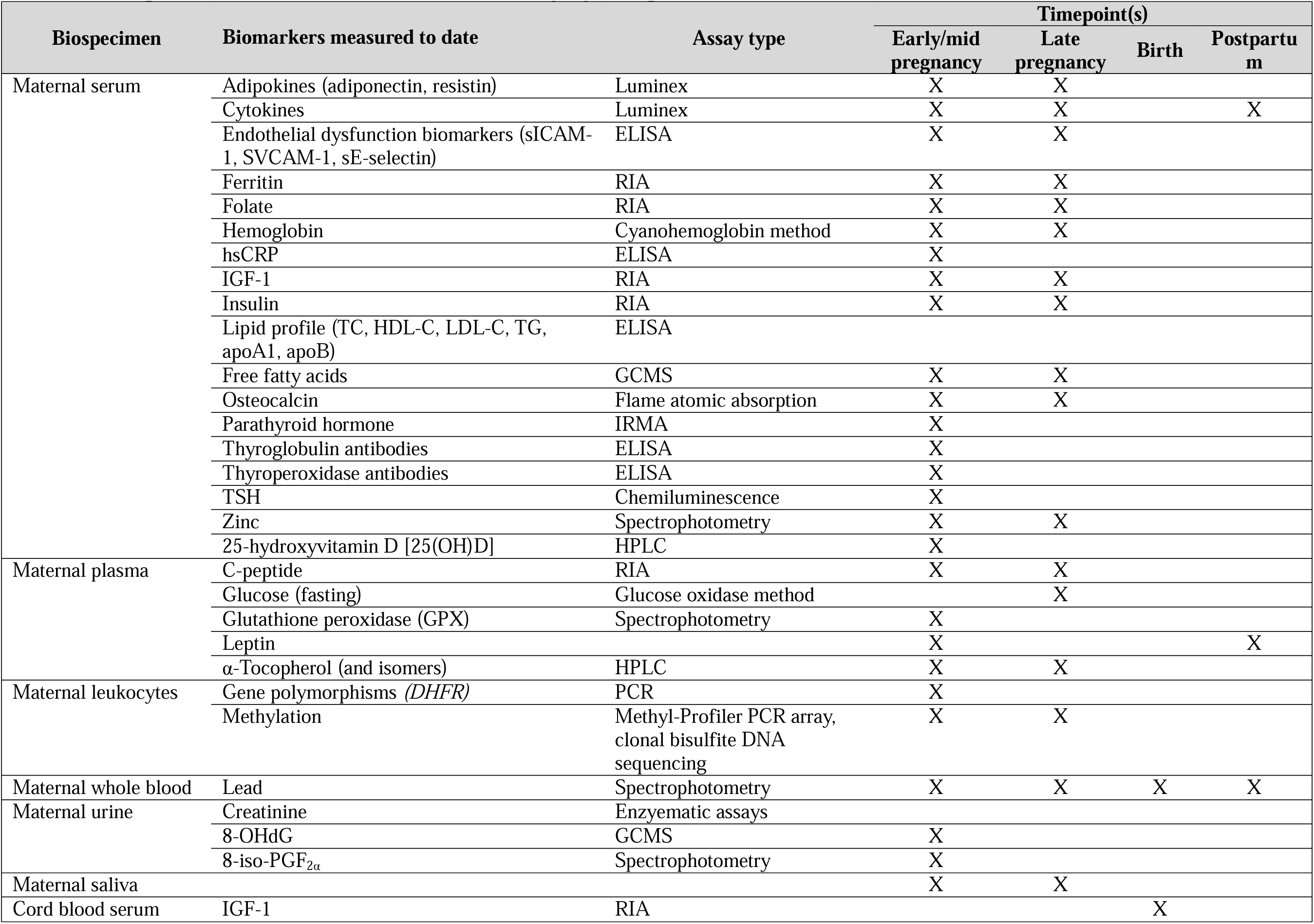
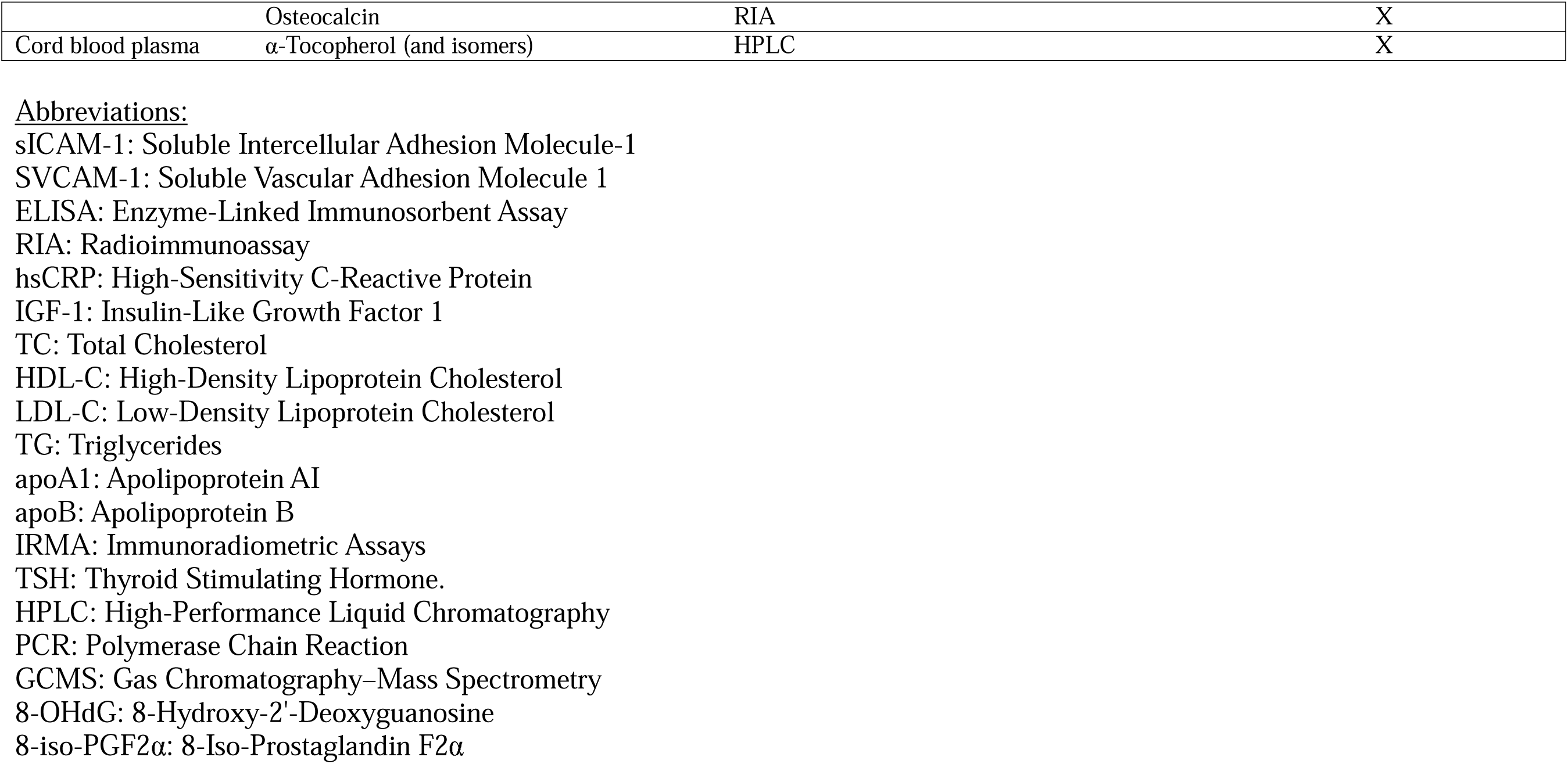
Biospecimens collected in the Camden Study by timepoint and biomarkers measured to date.

### Postnatal visits

Participants completed postpartum study visits approximately 4-6 weeks after delivery and up to 6 months postpartum (n=∼600 participants). These visits again included interviews with study staff. Participants reported on the extent and frequency of lactation, use of formula, and juice/sweetened beverages for infants, use of WIC, and any maternal supplementation (e.g., vitamins, iron, bone meal). Participants again underwent extensive anthropometry and provided biospecimens as described above.

## Findings to date

From 1985-2006, 4765 pregnant participants aged 12 years and older were recruited. The cohort reflects a population highly under-represented in perinatal cohort studies, with 45% Hispanic, 38% non-Hispanic Black, and 17% White participants and 98% using Medicaid in pregnancy. Baseline characteristics of the cohort are shown in **Table 3**. Below we summarize published Camden Study results across several key areas of focus.

**Table 3.** Descriptive statistics of mother-child dyads from the Camden Study (n=4765).

| **Maternal Characteristics** | **Mean±SD**<br>**or N (%)** |
| --- | --- |
| Age (years) | 20.37 ± 4.92 |
| Race/ethnicity |  |
| Hispanic | 1900 (39.87) |
| Black | 2217 (46.53) |
| Non-Hispanic White and<br>Other | 648 (13.60) |
| Nulliparous | 2143 (44.97) |
| Pre-pregnancy BMI (kg/m<sup>2</sup>) | 24.75 ± 5.81 |
| Obese (BMI ≥30 kg/m<sup>2</sup>) | 735 (15.65) |
| Smoking during pregnancy<br>(any) | 1033 (21.68) |
| Preeclampsia | 382 (8.01) |
| **Infant/Delivery**<br>**Characteristics** |  |
| Gestational age at delivery<br>(weeks) | 38.62 ± 3.06 |
| Preterm birth | 637 (13.36) |
| Spontaneous preterm | 528 (11.00) |
| Birth weight (g) | 3162 ± 586 |
| Low birth weight (<2500g) | 421 (9.47) |

### Nutrition, growth, and outcomes in adolescent pregnancies

One of the original primary goals of the Camden Study was to study the adolescent metabolic *milieu* (*i.e.*, transient higher levels of fasting insulin, elevations in growth hormone and insulin-like growth factors) in the context of pregnancy. Furthermore, the study aimed to ascertain whether: (1) adolescent growth and metabolism during pregnancy relates to maternal health; and (2) observed changes in body composition mediate the association between maternal growth and reduced infant birth weight.^31^ Linear growth in pregnancy was assessed using the KHMD, which measures a segment of the body less susceptible to the effects of lordosis and weight gain. We found pregnant adolescents have significant positive increments in knee height growth compared with small decrements in knee height exhibited by an adult pregnant comparison group indicating continued linear growth during pregnancy in adolesecents.^32–34^ Maternal linear growth in pregnant adolescents, furthermore, was associated with greater gestational weight gain and increased fat stores in the postpartum period at caloric intakes comparable with pregnant older controls.^35^ In addition, mothers who gained upper arm fat late in pregnancy or continued to accrue fat in the postpartum period had the largest gestational weight gains, had infants who were smaller at birth, and retained the most weight postpartum.^29^ Pregnant adolescents who were growing had infants with lower birth weights compared to infants of nongrowing adolescents and those of older pregnant controls.^25^ In testing potential mechanisms for these findings, maternal growth was found to be associated with a higher systolic/diastolic ratio detected by Doppler ultrasound, which may indicate impaired blood flow and has been associated with diminished fetal growth and other adverse perinatal outcomes. Leptin was also assessed as a potential biomarker for continued growth; findings showed that growing adolescents had higher leptin concentrations in gestation and postpartum than non-growing pregnant adolescents and older pregnant controls, and that a leptin surge by week 28 was associated with increased maternal growth and decreased birth weight.^26^

### Nutrition biomarkers and pregnancy outcomes

The Camden Study has provided valuable evidence on the critical role of nutrient intake and status in shaping birth outcomes, particularly focusing on preterm delivery and birth weight. Several published studies revealed that low levels of folate^36^ or iron^37^ and low-normal levels of selenium (lowest decile) during pregnancy^38^ were associated with increased risk of preterm delivery and/or low birth weight. Notably, the study was the first to investigate the relationship between dietary zinc intake and preterm delivery,^24^ as well as the first prospective study to report an association between folate intake and preterm delivery.^36^ A low intake of dietary zinc early in pregnancy was linked to an over threefold increase in the risk of very preterm delivery (before 33 completed weeks),^24^ while women with a low daily folate intake (less than 240 g/d) had approximately twice the risk of preterm delivery and infant low birth weight^36^ even after controlling for maternal characteristics, energy intake, and correlated nutrients. Lower concentrations of serum folate at week 28 were also linked to an increased risk of preterm delivery and low birth weight.^36^ Other vitamins such as Vitamin D and Vitamin E were also investigated in the cohort. Higher circulating concentrations of alpha-tocopherol were positively associated with various indicators of fetal growth, including increased birth weight for gestation and a reduced risk of infants being small for gestational age.^39^ With respect to Vitamin D, vitamin D insufficiency was linked to a two-fold increased risk of cesarean for prolonged labor,^40^ whereas high vitamin D intake was associated with increased infant size at birth.^41^ Secondary hyperparathyroidism, indicating vitamin D deficiency and impaired calcium metabolism, was also linked to over a two-fold risk of preeclampsia, and a two- to three-fold increased risk of small-for-gestational-age births and significantly lower birth weight, length, and head circumference.^42,43^

Consistent with the findings from individual studies looking at vitamins and minerals, study participants who started taking supplements in the first or second trimester experienced a two-fold reduction in the risk of preterm delivery, and for first trimester users, the risk of very preterm delivery was reduced more than fourfold.^44^ Prenatal supplement use was also associated with a reduced risk of infant low birth weight and very low birth weight.^44^ Regarding macronutrient intake, the women in the Camden study had a diet that was notably high in sugar, with approximately half of the carbohydrates in the diet attributed to sugar.^45,46^ High-sugar diet in this cohort was in turn linked to increased risk for delivering small-for-gestational-age infants.^45,46^

### Glucose regulation in pregnancy and risk factors for GDM

Markers and indices of glucose metabolism were measured in blood collected in the second and third trimester, as well as glucose levels following a 50-g glucose load administered between 24-28 weeks to screen for GDM. Most analyses within the Camden Study excluded women who met the 2000 American Diabetes Association (ADA) criteria for GDM (n=81).^47^ At the time of study recruitment and analysis, most clinical practices in the US utilized a two-step approach to diagnosing GDM, with a glucose threshold of >130 mg/dl following a 50-g glucose challenge. Many of the analyses within the Camden Study reported positive associations of what was considered to be sub-clinical elevated glucose levels with adverse perinatal outcomes^48^ and larger neonatal size.^48,49^ However, the ADA now supports a one-step criteria with a lower glucose threshold, which if applied to published work from the Camden Study would result in previously normoglycemic pregnancies being categorized as having GDM.^50^

Among participants without GDM, the dietary glycemic index calculated from recalls at 15, 20, 28 gestational weeks was positively correlated with glycosylated hemoglobin measured at GDM screening and with neonatal size.^51^ Estimated insulin resistance calculated by the homeostatic model of insulin resistance (HOMA-IR) in the second trimester was associated with greater gestational weight gain and postpartum weight retention.^52^ While there were no differences in fasting glucose levels between self-identified races/ethnicities, Black women had higher post-challenge glucose levels and greater insulin resistance.^53^

Several non-glycemic biochemical markers were measured including free fatty acids (FFAs) (e.g., palmitoleic, oleic, linolenic, myristic acids), inflammatory markers (c-peptide), adipokines/cytokines, and markers of amino acid metabolism (glutathione). Specific FFAs (palmitoleic, oleic, linolenic, myristic acids) were negatively associated with HOMA-IR and C-peptide, and palmitic, stearic, arachidonic, dihomo-γ-linolenic (DGLA) and docosahexaenoic acids were positively associated with HOMA-IR and C-peptide,^54^ with a higher total FAs observed among women with GDM or impaired glucose tolerance.^55^ Glutathione, a marker of amino acid metabolism, was positively associated with fat intake and fasting insulin, glucose, C-peptide, and insulin resistance.^56^

Elevated FFA levels measured on average at 30 weeks gestation were also found to be associated with a three-fold increased risk of preterm delivery, independent of pre-pregnancy obesity and other known risk factors for preterm delivery, including cigarette smoking, ethnicity, and prior preterm delivery.^57^

### Hormones, inflammatory biomarkers, and measures of oxidative stress

The Camden Study has explored a number of hormones, inflammatory biomarkers, and measures of oxidative stress as they relate to pregnancy outcomes. Pregnant women with elevated plasma leptin concentrations at baseline were predisposed to greater weight gains during pregnancy and more weight retention postpartum.^58^ Lower adiponectin levels were found to be associated with an increased risk of GDM and preeclampsia. Lower adiponectin levels were also associated with a higher risk of preterm delivery among Black women, but not Hispanic or White women.^59^ In addition, elevated levels of biomarkers of endothelial function linked to cardiovascular disease risk, including soluble intercellular adhesion molecule-1 (sICAM-1) and vascular cell adhesion molecule-1 (sVCAM-1), were associated with preterm delivery.^60^

We showed that high maternal oxidative stress, measured using urinary excretion of 8-iso-PGDF_2D_(isoprostane) and 8-oxo-7.8 dihydro-2-deoxyguanosine (8-OHdG), and captured in early pregnancy, was associated with several pregnancy outcomes: nearly five times higher odds of preeclampsia, shortened gestational duration, lower infant birth weight, and a distortion in the sex ratio at birth (57% males vs. 40% females).^61^ Lower 8-OHdG was also associated with the occurrence of minor congenital disabilities. Finally, the highest tertiles of high-sensitivity C-reactive protein, measured at entry and 28 weeks gestation, were associated with increased odds of early preterm delivery and pregnancy-induced hypertension among women with BMI <25 kg/m^2^.^62^

### Racial/ethnic disparities in biomarkers in pregnancy

Exposure to systematic and structural racism imparts undue physiological chronic stress, which we now understand to induce physiological “weathering” even in reproductive age women.^63^ In light of this, the Camden Study was one of the earliest studies to document racial/ethnic health disparities in perinatal complications and study the biochemical pathways that might underlie these disparities. First, we hypothesized that we might observe racial/ethnic differences in maternal lipids such as high density lipoprotein (HDL) that would lead to risk of preterm birth; however, no differences were observed.^64^ Second, we examined whether there were racial/ethnic differences in maternal adipokines during pregnancy. We reported that Black women had lower concentrations of adiponectin, a strong anti-inflammatory marker, as well as higher concentrations of resistin, a marker for inflammation and insulin resistance.^28^ The association between lower adiponectin levels and preterm birth was significant only in Black women and results persisted after adjustment for multiple confounders including BMI.^28,59^ Third, we examined racial/ethnic differences in glucose homeostasis in non-diabetic pregnant women, finding that Black women had significantly reduced insulin production and greater insulin resistance.^53^ These changes were stronger in the third trimester compared to the second.

## Strengths and limitations

The Camden Study has several important strengths that contribute to its distinct research value. First and foremost, the study recruited a unique population of pregnant women from a small number of clinics situated within an underserved urban area, bringing attention to a population that has been less represented in research. Of particular note was the recruitment of pregnant adolescents ages 13-18 years, a hard-to-reach population that is traditionally difficult to involve in research studies. The study had high rates of follow up, retaining > 90% of participants across pregnancy and collecting different biospecimens in each visit. Importantly, although this is a historical cohort recruited in the 1990-2000s, the disparities in the rates of adverse perinatal outcomes are still prevalent^65^ reinforcing its continued relevance. There are unique opportunities to link the Camden Study to existing administrative data and registries (e.g., to environmental monitoring data) and the low mobility of this population may also provide opportunities to re-engage participants, who would now be in their 30s-70s, and their young adult children.

The large sample size and high representation of minority groups (85% of the cohort) in the Camden Study has allowed for the assessment of health disparities, finding evidence of differential risk of health outcomes by race/ethnic group. For example, we observed that Black women had a more favorable lipid profile compared to non-Hispanic White women, which may protect against adverse pregnancy outcomes, yet they still had a higher risk of fetal growth restriction and preterm birth. Today, we understand race/ethnicity to be a social construct that correlates with a variety of factors that are specific to social context including experiences of racism, discrimination, some aspects of culture, socioeconomic status, and many other factors that influence health such as diet and environmental pollution.^66,67^ In most of the Camden Study analyses, where differences were observed, they mirrored patterns of health disparities leading to different perinatal complication rates in the general population. Future analyses focused on social determinants of health could include stress biomarkers, environmental chemical exposures, and multiple social and health indexes.

A major strength of the Camden Study is its rich prospective data collected during pregnancy and biospecimen repository. In combination with previously assayed information (**Table 2**), the biospecimen repository has and will continue to help provide a detailed understanding of biological markers of perinatal health and support future research endeavors. The Camden Study is also one of the few pregnancy cohorts to include repeated and longitudinal assessments of diet in a racially/ethnically diverse population. Importantly, the availability of comprehensive anthropometric data (e.g., skinfolds, circumferences) at multiple timepoints during pregnancy enables the ability to understand dynamic changes in body composition in pregnant persons over time.

The Camden Study presents a distinct and valuable opportunity to investigate untreated GDM, due to use of a higher threshold for diagnostic criteria than what is currently accepted. This approach allows researchers to explore a subset of women who might not have been diagnosed with GDM by contemporary standards, yet many of these individuals would now receive a GDM diagnosis under current criteria. Future studies could provide a more nuanced understanding of the impact of maternal glycemia and its relationship to other biochemical markers impacted by insulin dynamics (e.g., adipokines, FFAs, amino acids), contributing to a more comprehensive and up-to-date assessment of its implications for both maternal and fetal well-being.^68^

At the same time, potential shifts in clinical guidelines over time may limit our ability to compare some findings to present-day standards (e.g. changes to preeclampsia diagnosis guidelines).^69^ This challenge, however, opens an avenue for reanalysis in the current context, providing an opportunity to contextualize and reinterpret results. The cohort’s focus on “healthy” pregnant women who started prenatal care at or before 17 weeks may additionally limit the generalizability to women who do not engage in prenatal care, which can occur more often in populations with low resources.^70^ Finally, limited data are available on other measures of interest including racism/discrimination, physical activity, and mental health diagnoses/behavioral health factors, though linkages with neighborhood-level factors remain a possibility.

## Conclusions

The Camden Study data and biorepositories are well-positioned to support future research aimed at better understanding perinatal health in under-represented women and infants. The integration of new high throughput assays for -omics studies, coupled with the integration of these -omics data to existing datasets using artificial intelligence, may be a particularly promising approach for enhancing prior Camden Study research. Importantly, Camden County was, and continues to be one of the most polluted areas in N.J. and the U.S. with 13 designated Superfund sites.^71,72^ These particular characteristics make the Camden Study pregnancy cohort a unique population to study environmental exposures and pregnancy/birth outcomes, which have received little attention in this cohort to date. While we acknowledge that the quality of long-term banked specimens may not be universally appropriate for all types of analyses, research has demonstrated the feasibility of conducting analyses on samples preserved at −80°C for extended periods.^73^ Finally, the strategic linkage of the Camden Study to various resources, including medical records through the Camden Cooperative and administrative datasets, as well as potential for recontact of participants 20-30 years after initial participation may provide key insights into trajectories of maternal and child health across the life course.

## Data Availability

Researchers interested in possible collaborations utilizing data and/or biospecimens from the "Camden Study" should contact the Principal Investigators (EB, ZRN). Interested collaborators will be requested to produce a concept proposal for their analysis, which the Camden Study Executive Committee will review. Following approval of the concept proposal, an analysis plan and proof of IRB approval must be submitted by collaborators to the Executive Committee before data/samples can be received. Collaboration requests will be deliberated on an ongoing basis.

## Consent for publication

Not applicable.

## Acknowledgements

We thank the Camden Study participants and the Camden Study research support staff as well as prior members of the Camden Study investigative team. We especially thank Theresa Scholl for her leadership on the Camden Study and review of the manuscript.

## Funding

The Camden Study was supported by historical grants from the National Institutes of Health (R01HD018269/Scholl; R01ES007437/Scholl; R01HD038329/Scholl; R21DK078865/Chen; R21HD058128/Scholl; R21HD061763/Chen; R01MD007828/Chen) with additional current support provided by P30ES005022.

## Competing interests

None declared.

## Authors’ Contributors

TS and XC originally designed and led the study. EB and ZRN now provide leadership on the study. EB, ZRN, and SS led manuscript development with intellectual content for subsections provided by all coauthors. All authors provided final approval for the version to be published and agree to be accountable for all aspects of the work.

## Ethics approval and consent to participate

This study was approved by the Institutional Review Board at Rutgers University.

## Availability of data and materials

Data are available upon reasonable request. Collaborations with the research team are welcome. Available data are listed in **Tables 1 and 2**. Researchers interested in collaboration are invited to contact the Principal Investigators (EB, ZRN). Data access requests will be assessed by the Executive Committee of the Camden Study.

## Footnotes

a *Nota bene*, as Camden Study investigators and members of the epidemiological scientific community, we are committed to using inclusive language, especially in relation to gender. We choose to use gendered terminology following the rationale for using female-sexed language in studies of maternal and child health. Additionally, all the prior studies from the Camden Study that are summarized herein used female terminology to describe their pregnancy cohort, as a pregnancy can only occur in individuals who are female at birth. In this article, we use the term ‘women’ throughout, but acknowledge that not all individuals who experienced a pregnancy may self-identify as a woman.

## Notes

### Competing Interest Statement

The authors have declared no competing interest.

### Author Declarations

The Institutional Review Board at Rutgers University gave ethical approval for this work.

